# Global burden of preterm birth among newborns from 1990 to 2023 and projections to 2050: a retrospective trend analysis and projection study

**DOI:** 10.64898/2026.03.21.26348954

**Authors:** Haixia Wan, Xiuming Zhong, Xiaoting Zhang

## Abstract

Based on the 2023 Global Burden of Disease (GBD) database, this study analyzed the global burden of preterm birth from 1990 to 2023 and predicted its development trend by 2050, while exploring the disparities in disease burden across regions with different Socio-demographic Index (SDI) levels, income groups and countries. A retrospective trend analysis was conducted to collect data on preterm birth incidence, prevalence, death and disability-adjusted life years (DALYs) in 204 countries and regions worldwide from 1990 to 2023 from the GBD 2023 database. ARIMA model (p=2,d=1,q=1) and grey prediction model (GM(1,1)) were combined to predict the preterm birth burden from 2023 to 2050. In 2023, preterm birth was the primary cause of the global neonatal disease burden, with its four core indicators significantly higher than other neonatal diseases. From 1990 to 2023, the global incidence, death and DALYs of preterm birth decreased to 0.91, 0.44 and 0.52 times of the 1990 levels respectively, while the prevalence increased to 1.54 times of the baseline. Projection results showed that by 2050, the incidence, death and DALYs of preterm birth would drop to 0.79, 0.08 and 0.32 times of the 2023 levels, and the prevalence would rise to 1.23 times of 2023. Low SDI regions, lower-middle income countries, as well as India and Nigeria, bore the heaviest disease burden. Over the past three decades, the global acute health burden of preterm birth such as death has decreased notably, but the continuous rise in prevalence and severe regional and age disparities remain prominent public health challenges. The 0-6 days and 6-11 months age groups are the key time windows for preterm birth intervention. It is urgent to implement targeted prevention and control measures for low SDI regions and lower-middle income countries to reduce the global burden of preterm birth.

## Introducation

Neonatal health is a crucial component of global public health, and is closely associated with the achievement of the United Nations Sustainable Development Goals (SDGs) and the promotion of universal health coverage^1^. Preterm birth, defined as live birth before 37 completed weeks of gestation, is the leading cause of death in neonates and children under 5 years old worldwide^2, 3^. According to the World Health Organization (WHO) statistics in 2023, about 13.4 million infants were born prematurely globally in 2020, accounting for approximately one-tenth of all live births^2^. In addition to the high acute death risk, preterm birth is also associated with a series of long-term adverse outcomes such as cerebral palsy, neurodevelopmental delay and chronic respiratory diseases^4-6^, which not only bring heavy health burdens to individuals, but also cause considerable socioeconomic pressure to families and societies^7-9^.

In recent years, global initiatives such as the WHO Every Newborn Action Plan launched in 2014 and the SDG 3 target of ending preventable newborn deaths by 2030 have effectively promoted the development of maternal and neonatal health services in various regions, and improved the diagnosis and treatment capacity of preterm birth to a certain extent^1, 10^. However, existing studies have shown that there are significant disparities in the global burden of preterm birth across regions with different SDI levels and income groups, with low and middle income countries bearing more than 90% of the global disease burden^5, 11^. Although some studies have analyzed the trend of preterm birth burden based on GBD data, most of them adopted pre-2023 database data, lacked reliable long-term projection models up to 2050, and failed to fully explore the age-stratified characteristics of preterm birth burden and the internal relationship between the decline in death and the rise in prevalence. In addition, the key intervention time windows for different age groups and the targeted prevention and control strategies for high-burden regions are still not clear.

The 2023 update of the GBD database provides standardized and comprehensive disease burden data for 204 countries and regions worldwide, which lays a solid foundation for an accurate assessment of the global preterm birth burden. Based on the GBD 2023 database, this study conducted a retrospective analysis of the global preterm birth burden from 1990 to 2023, and combined multiple prediction models to forecast its development trend by 2050. Meanwhile, this study explored the disparities in preterm birth burden across SDI levels, World Bank income groups and major countries, and identified the key age-specific intervention time windows and modifiable risk factors. It is expected to fill the research gaps in the current global preterm birth burden research, and provide a scientific basis for formulating evidence-based and stratified preterm birth prevention and control strategies, so as to promote the equitable development of global maternal and neonatal health services.

## Methods

### Data sources

GBD 2023 is a large-scale international research initiative that integrates data from diverse sources, including population censuses, surveys, vital statistics, disease registration systems, and environmental monitoring data. Following rigorous evaluation and standardization, these data are accessible via the data access tool (http://ghdx.healthdata.org/gbd-results-tool), with the overarching aim of assessing the global disease burden. Since this study involves no identifiable personal information, participants and the general public were not involved in data collection, nor did they provide input on research questions, outcome measures, or study design. Ethical approval and informed consent were therefore deemed unnecessary and impracticable. GBD 2023 analyzed the disease burden across 204 countries and regions at the global, regional, and national levels, and stratified the study populations into four categories by the SDI: low SDI, low-middle SDI, middle SDI, and high-middle SDI. The study also used the World Bank income classification to divide the populations into four income groups: low income, lower-middle income, upper-middle income, and high income regions.

### Definitions in GBD 2023

The GBD Study provides estimates of incidence, prevalence, DALYs, deaths, and those attributable to specific risk factors. Incidence refers to new cases in a population over a period, while prevalence is the proportion of a population affected by a disease at a specific time. From 1990 to 2023, global neonatal disease burden across all age groups was assessed using death and DALYs. Death rate, a key public health indicator, reflects the proportion of deaths in a population and is used to evaluate disease severity and healthcare effectiveness. DALYs combine years of life lost (YLLs) and years lived with disability (YLDs). The GBD Study assesses risk factors’ impact on DALYs by estimating disease burden changes at the theoretical minimum risk level. The 2023 GBD Diseases and Injuries Collaborators provided a comprehensive overview, with 500 replicate simulations to generate sample-level estimates and 95% uncertainty intervals (UIs). Visualized and tabular data are available at https://vizhub.healthdata.org/gbd-comparison/ and https://vizhub.healthdata.org/gbd-results/, respectively. All analyses followed GBD protocols and guidelines strictly.

### Risk Factors in GBD 2023

The GBD Study categorizes all risk factors into a four-tier hierarchical framework and provides a comprehensive summary of them. The first tier divides risk factors into four major categories: environmental, occupational, behavioral, and metabolic. Detailed information on these four categories is available in the GBD 2023 online resources (https://ghdx.healthdata.org/gbd-2021/sources?components.6&risks.367&location).

This study explores the key contributions of these risk factors to disease risk across all age groups and the global disease burden, and assesses their specific impacts on total deaths, DALYs, as well as disease prevalence and incidence.

### Data Prediction and Analysis

Statistical analyses were performed with IBM SPSS Statistics 26.0 software. Age-standardized rates of all indicators were calculated based on the World Standard Population. For the projection of preterm birth burden from 2024 to 2050, a combined prediction approach integrating the ARIMA model (p=2, d=1, q=1) and grey prediction model GM(1,1) was adopted, with all models meeting the criteria of good fit (R²>0.85) and passing the residual white noise test (P>0.05). The Cochran-Armitage trend test was applied to evaluate the statistical significance of temporal changes in global preterm birth burden indicators, with a two-tailed P value < 0.05 defined as statistically significant.

### Data Visualization

Data visualization was performed using GraphPad Prism 9.0 software, which accurately presented the data analysis results of all experiments. For example, bar charts could intuitively display the indicator differences among different experimental groups, facilitating quick comparison of data levels; line charts could dynamically reflect the changing trends of target variables with the passage of time or other intervention factors, clearly showing the fluctuation rules of data. In the process of chart production, this study uniformly standardized the format of elements such as color matching, font size, and axis label styles to enhance the identification and standardization of charts, further improve their readability and visual aesthetics, and ensure that data presentation is both scientific and rigorous, as well as intuitive and easy to understand.

## Results

### Preterm birth as the leading cause of neonatal disease burden in 2023

According to the GBD 2023 database statistics, the global incidence of neonatal diseases in 2023 was 26.1 million (95%UI: 25.9-26.3), the prevalence was 159.8 million (95%UI: 144.9-174.7), the death was 1.6 million (95%UI: 1.5-1.7), and the DALYs were 170.0 million (95%UI: 158.6-183.0). The incidence of preterm birth was 21.3 million (95%UI: 20.9-21.2), which was 5.88 to 54.36 times that of other neonatal disease etiologies. The prevalence was 125.5 million (95%UI: 108.9-142.5), 6.42 to 22.38 times that of other etiologies. The death was 0.6 million (95%UI: 0.5-0.7), 1.11 to 20.55 times that of other etiologies. The DALYs were 70.2 million (95%UI: 60.6-81.4), 1.28 to 15.03 times that of other etiologies (Figure 1).

**Figure 1.**
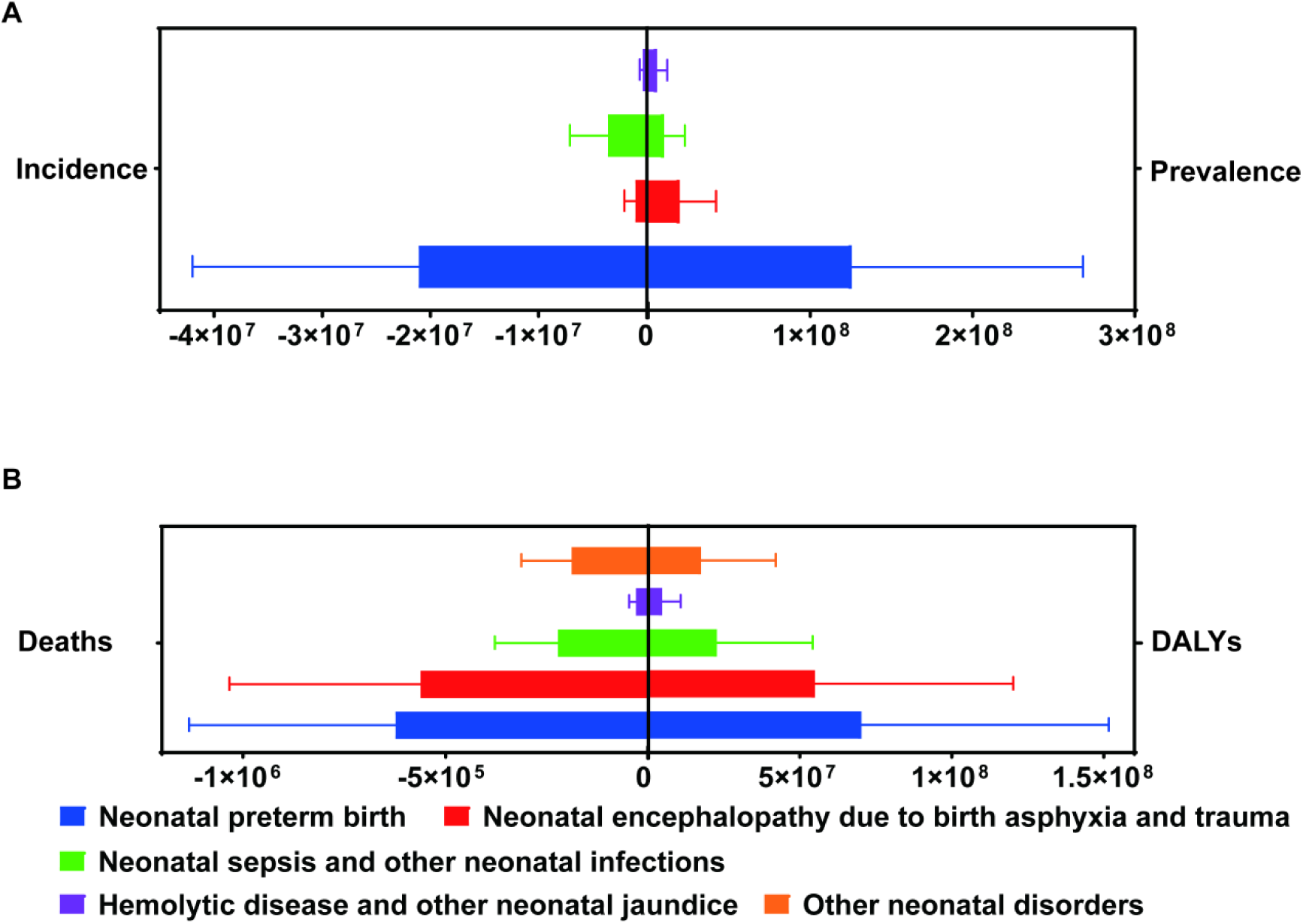
Preterm Birth, Neonatal Encephalopathy Due to Birth Asphyxia and Trauma, Neonatal Sepsis, and Other Neonatal Diseases Worldwide in 2023.

### Global preterm birth burden from 1990 to 2023 and projections to 2050

The incidence decreased from 23.0 million (95%UI: 22.9-23.2) in 1990 to 21.1 million (95%UI: 20.9-21.2) in 2023, a decrease to 0.91 times of the baseline. The death dropped from 1.4 million (95%UI: 1.2-1.7) to 0.6 million (95%UI: 0.5-0.7), a decrease to 0.44 times of 1990. The DALYs decreased from 135.3 million (95%UI: 117.9-156.9) to 70.2 million (95%UI: 60.6-81.4), a decrease to 0.52 times of the initial level. In contrast, the prevalence of preterm birth increased continuously from 81.5 million (95%UI: 70.0-91.6) in 1990 to 125.5 million (95%UI: 108.9-142.5) in 2023, an increase to 1.54 times of the 1990 level. In 2050, the projected incidence was 16.6 million (95%UI: -2.8-36.0). The prevalence was 154.5 million (95%UI: 128.0-181.0). The death was 49,058.14 (95%UI: -0.8-0.9 million). The DALYs were 22.7 million (95%UI: -58.0-103.5). Compared with the 2023 levels, the incidence, death and DALYs would decrease to 0.79, 0.08 and 0.32 times respectively, while the prevalence would increase to 1.23 times (Figure 2).

**Figure 2.**
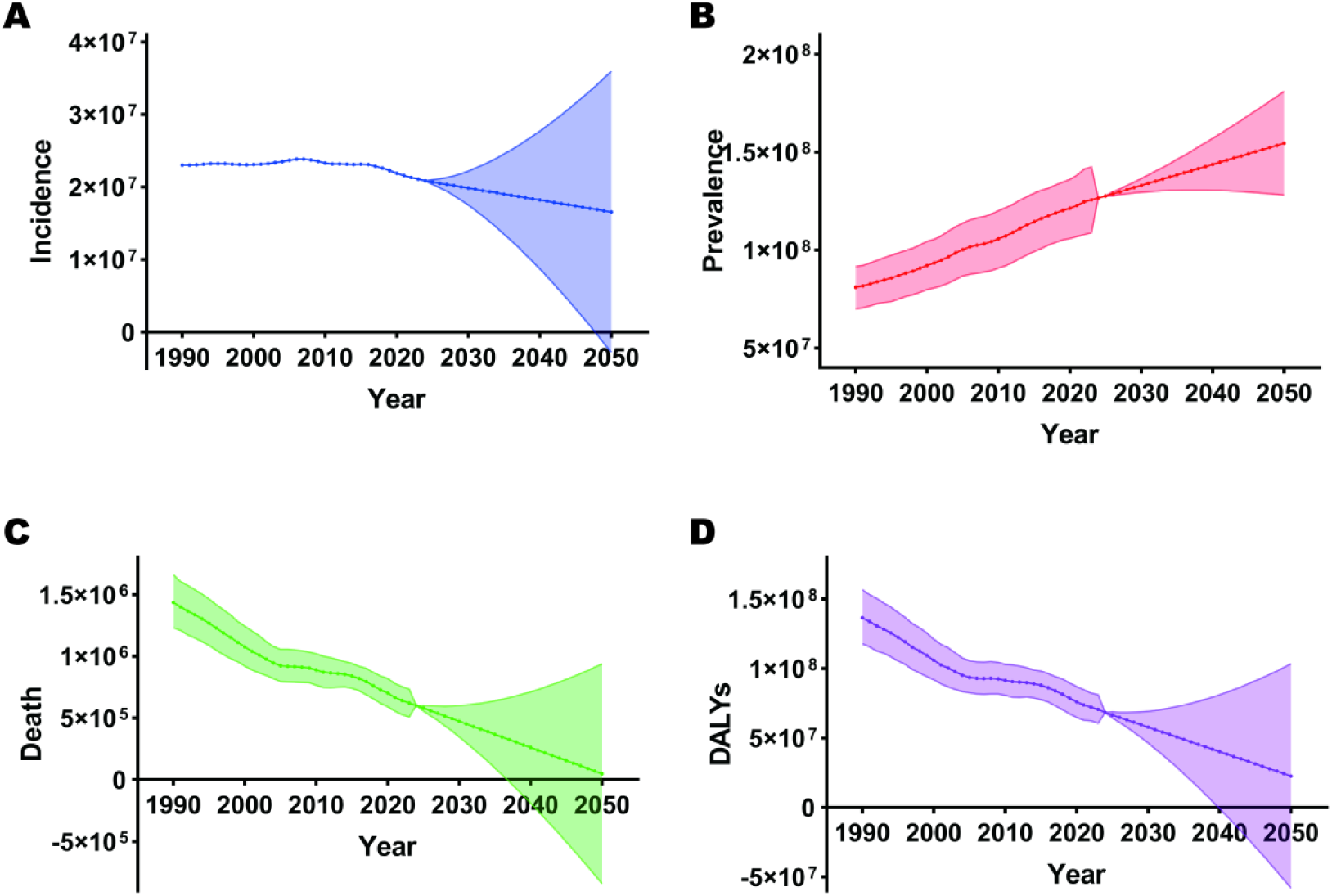
Incidence (A), Prevalence (B), Deaths (C), and DALYs (D) of trends in preterm birth among neonates from 1990 to 2050.

### SDI-stratified preterm birth burden in 2023

Among infants under 1 year old, the incidence of preterm birth was the highest in low SDI regions (9.5 million (95%UI: 9.4-9.6)), and the lowest in middle SDI regions (2.1 million (95%UI: 2.1-2.2)). In the age groups of 0-6 days, 7-27 days, 1-5 months, 6-11 months, 12-23 months and 2-4 years, the prevalence, death and DALYs of preterm birth were all the highest in low SDI regions. Among them, the 6-11 months age group had the highest prevalence (4.0 million (95%UI: 3.8-4.2)) in low SDI regions. Middle SDI regions had the lowest indicators in all age groups, with the 0-6 days age group having the lowest prevalence (41,721.20 (95%UI: 40,686.65-42,568.91)). In all six age groups, the 0-6 days age group had the highest death due to preterm birth (0.2 million (95%UI: 0.2-0.3)) in low SDI regions. The lowest death was found in middle SDI regions for the 7-27 days and 1-5 months age groups, and in high-middle SDI regions for the other age groups. The lowest death was observed in the 2-4 years age group in high-middle SDI regions (86.75 (95%UI: 69.28-106.28)). The 0-6 days age group in low SDI regions also had the highest DALYs (23.2 million (95%UI: 18.5-28.2)). The lowest DALYs were found in high-middle SDI regions for the 0-6 days and 6-11 months age groups, and in middle SDI regions for the other age groups. The lowest DALYs were in the 12-23 months age group in middle SDI regions (48,008.19 (95%UI: 39,054.72-60,171.77)) (Figure 3).

**Figure 3.**
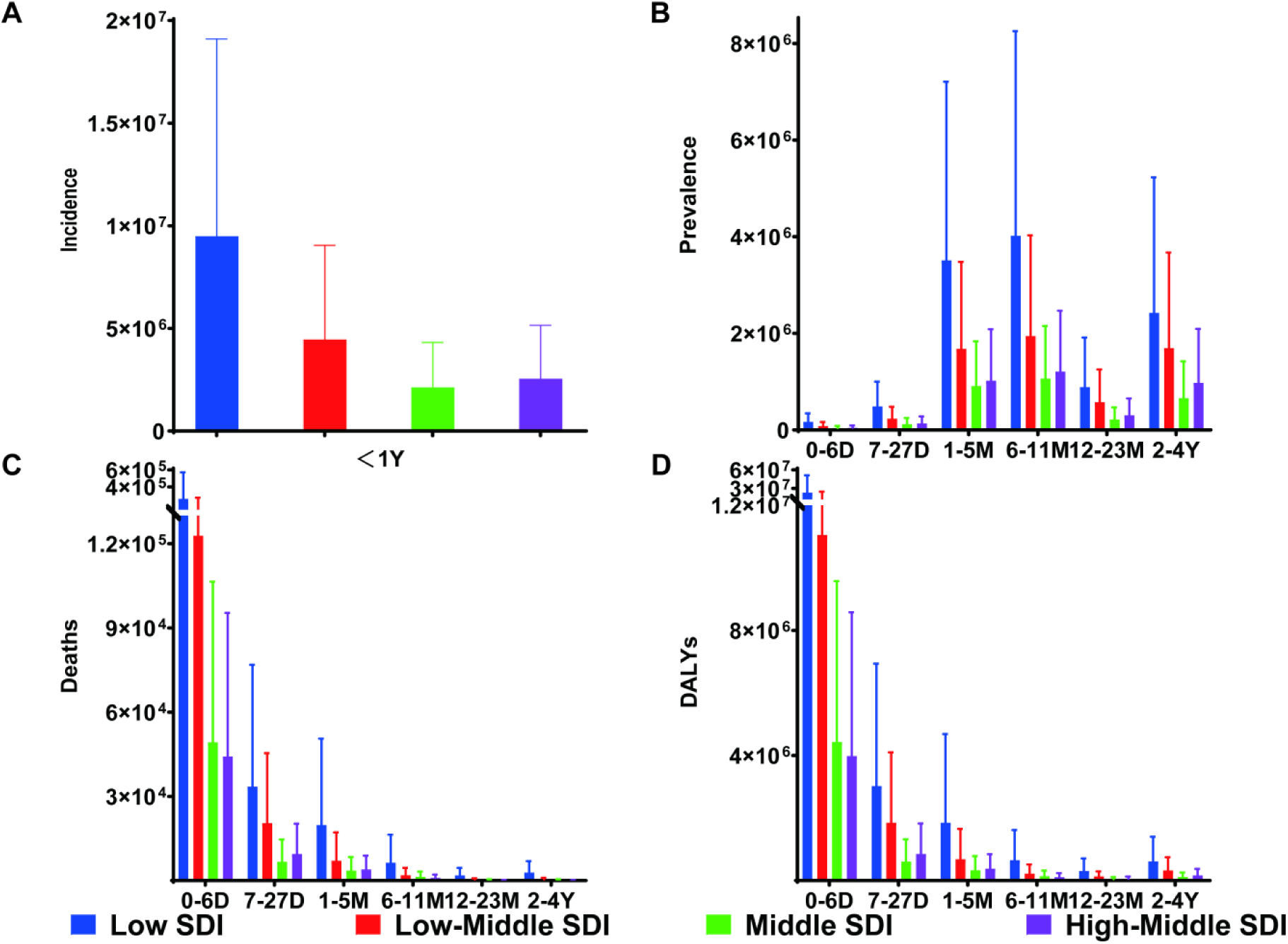
Incidence (A), Prevalence (B), Deaths (C), and DALYs (D) of Age-Stratified SDI Regions in 2023.

### World Bank income-stratified preterm birth burden in 2023

Across all age groups, the incidence, prevalence, death and DALYs of preterm birth were all the highest in lower-middle income regions, and the lowest in high-income regions. In the age groups of 0-6 days, 7-27 days, 1-5 months, 6-11 months, 12-23 months and 2-4 years, the 6-11 months age group in lower-middle income regions had the highest prevalence (5.6 million (95%UI: 5.2-5.9)), and the 0-6 days age group had the lowest prevalence (28.2 million (95%UI: 27.8-28.7)). The 0-6 days age group had the highest death and DALYs in all income groups: the death in lower-middle income regions was 0.3 million (95%UI: 0.3-0.4), and the DALYs were 29.1 million (95%UI: 24.0-34.5). The lowest death was found in the 2-4 years age group in high-income regions (38.95 (95%UI: 34.65-43.53)), and the lowest DALYs were in the 12-23 months age group in high-income regions (23,039.23 (95%UI: 18,169.74-28,246.62)) (Figure 4).

**Figure 4.**
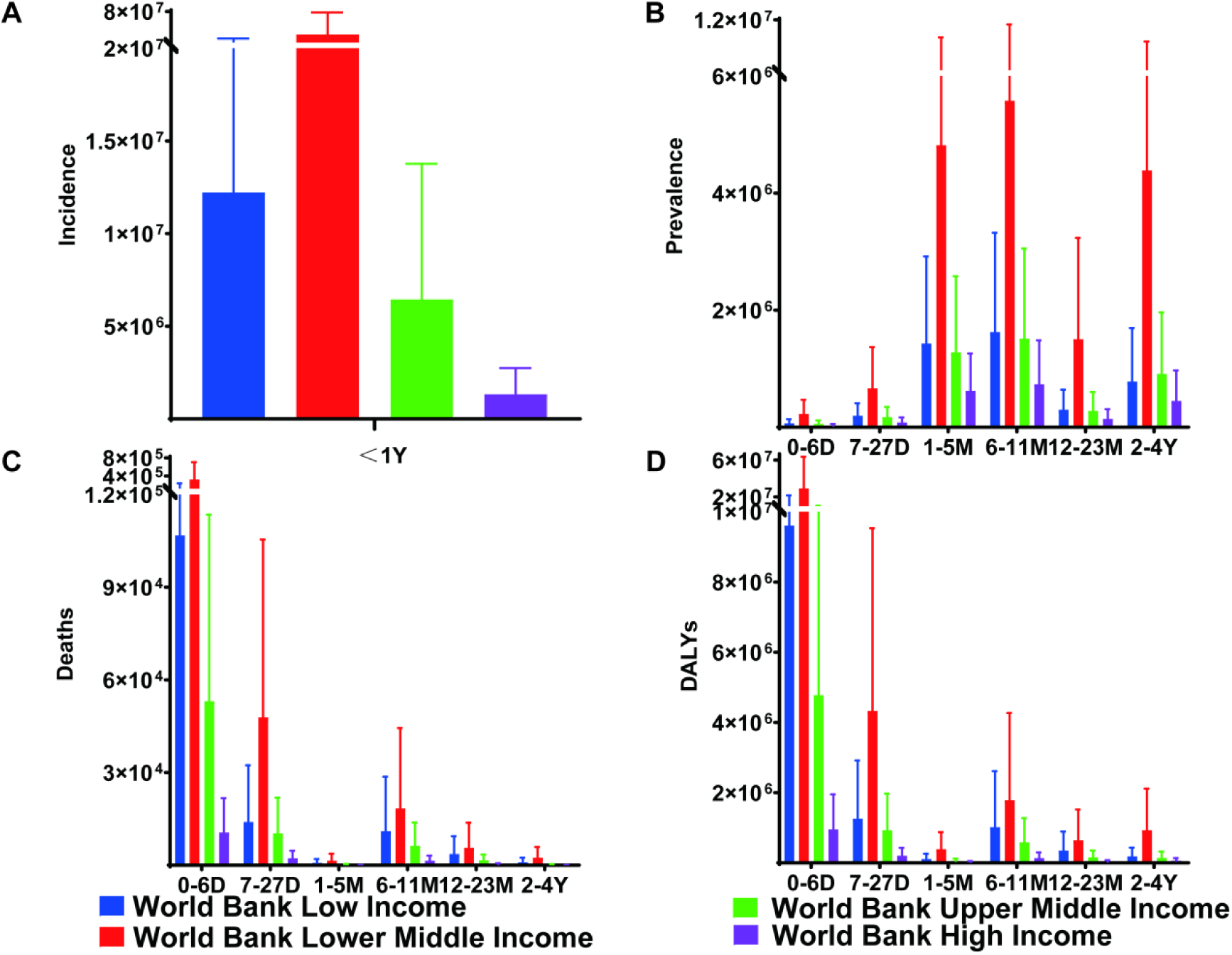
Incidence (A), Prevalence (B), Deaths (C), and DALYs (D) by Age-Stratified World Bank Income Regions in 2023.

### Country-stratified preterm birth burden in 2023

Across all age groups, India had the highest incidence, prevalence, death and DALYs of preterm birth. The only exception was that Nigeria had the highest death in the 6-11 months, 12-23 months and 2-4 years age groups. Brazil had the lowest incidence of preterm birth among infants under 1 year old. In the six age groups, the 6-11 months age group in India had the highest prevalence (2.6 million (95%UI: 2.4-2.8)). The lowest prevalence was found in Central Europe, Eastern Europe, and Central Asia for the 12-23 months age group, in Indonesia for the 2-4 years age group, and in Brazil for the other age group. The lowest prevalence was in the 0-6 days age group in Brazil (7.6 million (95%UI: 7.4-7.7)). The 0-6 days age group in India had the highest death (0.1 million (95%UI: 0.1-0.2)). The lowest death was found in Indonesia for the 1-5 months, 6-11 months and 12-23 months age groups, and in Brazil for the other age groups. The lowest death was in the 12-23 months age group in Indonesia. India also had the highest DALYs in the 0-6 days age group (11.6 million (95%UI: 9.5-13.9)). The lowest DALYs were found in Central Europe, Eastern Europe, and Central Asia for the 12-23 months and 2-4 years age groups, in Indonesia for the 1-5 months and 6-11 months age groups, and in Brazil for the other age groups. The lowest DALYs were in the 12-23 months age group in Central Europe, Eastern Europe, and Central Asia (5,841.23 (95%UI: 4,476.77-7,296.36)) (Figure 5).

**Figure 5.**
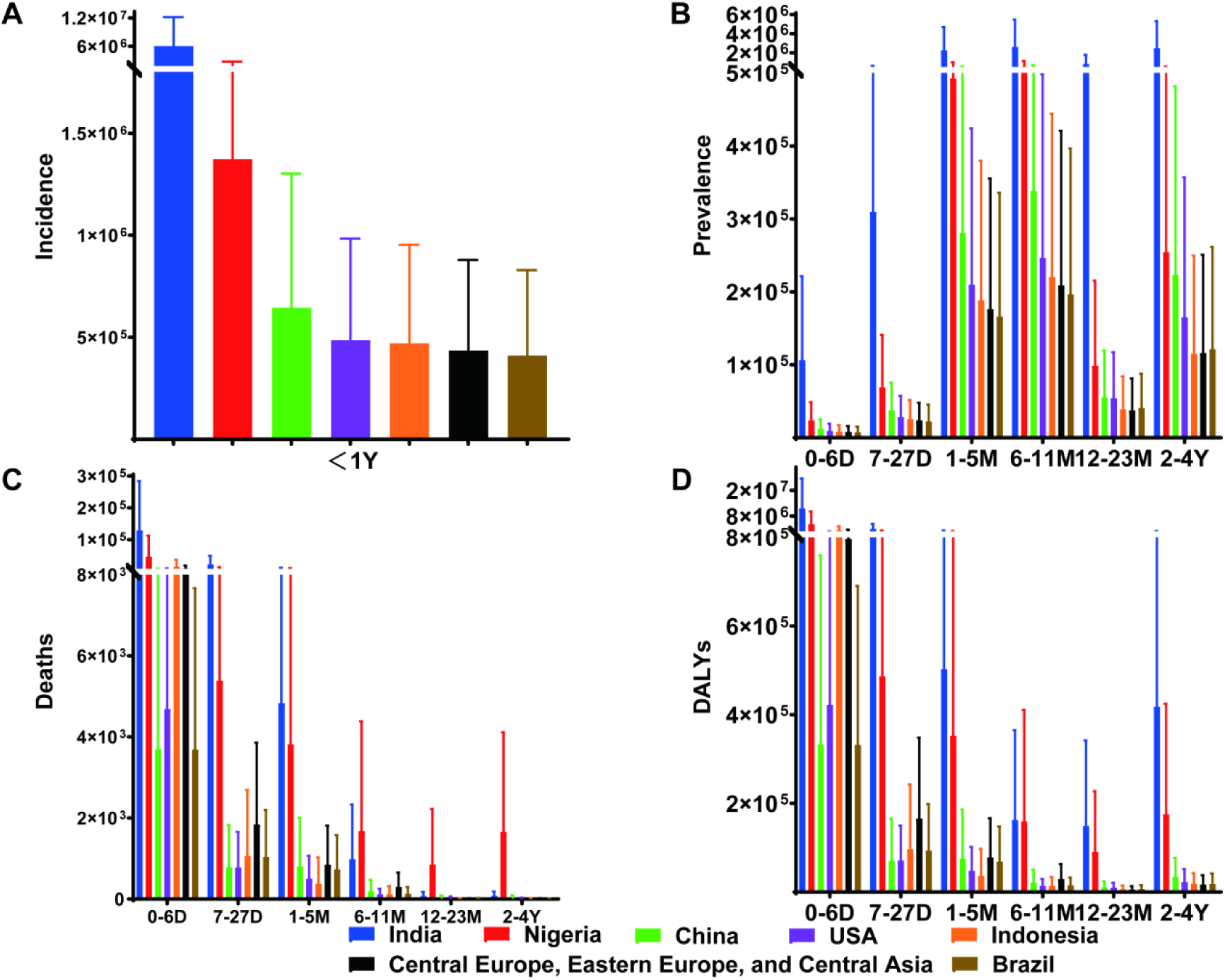
Incidence (A), Prevalence (B), Deaths (C), and DALYs (D) by Age-Stratified Countries in 2023.

## Discussion

Based on the latest GBD 2023 database, this study completed the analysis of the global preterm birth burden from 1990 to 2023 and the projection by 2050, and conducted in-depth stratified analysis from the perspectives of SDI level, income group and country. The results confirm that preterm birth is still the primary cause of the global neonatal disease burden, and the identified temporal and regional disparities provide important empirical evidence for formulating targeted global preterm birth prevention and control strategies.

The projection results to 2050 show that the global preterm birth incidence, death and DALYs will continue to decline, and the prevalence will rise to 1.23 times of the 2023 level. This trend indicates that the global prevention and control of preterm birth will face two core tasks in the next 30 years. On the one hand, it is necessary to strengthen primary prevention to further reduce the incidence and death of preterm birth. On the other hand, it is urgent to carry out secondary and tertiary prevention to cope with the growing burden of long-term sequelae of preterm birth.

A notable difference between this study and previous studies (such as a 2021 study that projected a significant increase in global preterm birth incidence^12^) is that this study found that the global preterm birth incidence will continue to decline by 2050. The results of this study provide a more optimistic and evidence-based prospect for global preterm birth control, but at the same time, it also emphasizes that the persistent regional inequities and the continuous rise in prevalence are still urgent problems to be solved.

The significant decline in the global preterm birth incidence, death and DALYs over the past three decades fully reflects the effectiveness of global maternal and neonatal health intervention measures. The popularization of the WHO in “Every Newborn Action Plan”^10^, the promotion of SDG 3 targets^1^, and the improvement of basic prenatal care and neonatal resuscitation technology in low and middle income regions have all played a key role in reducing the acute burden of preterm birth^13^. However, the 1.23-fold increase in the global prevalence of preterm birth has revealed a prominent public health contradiction^11^. The advancement of neonatal intensive care technology has significantly improved the survival rate of preterm infants, but at the same time, a large number of preterm survivors are faced with long-term sequelae such as cerebral palsy, neurodevelopmental delay and respiratory dysfunction^3, 14^. It leads to the continuous rise of prevalence.

There is severe inequity in the global preterm birth burden, with low SDI regions, lower-middle income countries, India and Nigeria bearing the main disease burden. This regional disparity is the result of the combined action of multiple factors. The insufficient neonatal care resources in high-burden regions, such as the lack of neonatal intensive care units, professional medical staff and essential therapeutic drugs such as pulmonary surfactant, which leads to the low rescue success rate of preterm infants^13, 15, 16^. The high prevalence of modifiable maternal risk factors in these regions, such as malnutrition, low educational level, PM_2.5_ exposure and inadequate prenatal care, which increases the incidence of preterm birth. The poor socioeconomic conditions, such as poverty, gender inequality and imperfect health system governance, restrict the implementation of maternal and neonatal health intervention measures^17-21^. The age-stratified results further clarify the key intervention time windows of preterm birth. The 0-6 days perinatal period is the critical stage to reduce death, which requires the improvement of acute neonatal care capacity^22-24^. The 6-11 months period is the key stage to reduce the long-term disease burden, which needs to strengthen postnatal follow-up, nutritional support and early screening of developmental disabilities^25^.

Ambient and household PM_2.5_ exposure is still one of the main modifiable risk factors for preterm birth, especially in low SDI regions^20, 26^. Previous studies have shown that PM_2.5_ exposure is associated with reduced gestational age and low birth weight of newborns. The first and second trimesters of pregnancy are the most vulnerable periods^27, 28^. In clinical practice, this finding reminds medical staff to carry out prenatal air pollution exposure screening in high-risk regions. And it provides personalized health guidance for pregnant women, such as reducing outdoor activities during periods of high PM_2.5_ concentration and using air purifiers at home to reduce indoor PM_2.5_ exposure^27, 29-32^. At the public health level, strict environmental pollution control measures to reduce PM_2.5_ emissions are not only an important measure to protect the ecological environment^33^, but also a critical intervention to improve maternal and neonatal health. It can effectively reduce the incidence of preterm birth and the corresponding medical and health costs^34-36^.

Maternal malnutrition, especially the deficiency of micronutrients such as vitamin D, copper, zinc, selenium, manganese, and chromium also help reduce the burden of preterm birth^37-41^, is a key risk factor for preterm birth. The 6-11 months age group has the highest prevalence further emphasizes the importance of postnatal nutritional supplementation for preterm survivors. Clinical evidence shows that vitamin D supplementation for preterm infants (such as intramuscular injection of 10,000 IU/kg vitamin D_3_ for infants with gestational age <34 weeks) can significantly reduce the incidence of vitamin D deficiency and its associated complications such as metabolic bone disease and respiratory infections^42-46^. This study supports the implementation of routine micronutrient level screening for pregnant women and preterm infants in low SDI regions. Targeted supplementation based on screening results can significantly reduce preterm birth risk and improve the long-term prognosis of preterm infants.

The severe preterm birth burden in low SDI and lower-middle income regions is fundamentally due to the inadequate access to quality prenatal care and neonatal care^11, 17, 47-49^. For example, lack of timely progesterone supplementation and insufficient training in neonatal resuscitation are key avoidable causes of high preterm birth death^13, 50-52^. Strengthening the construction of primary health care systems is the most effective way to reduce the preterm birth burden in high-burden regions, including training front-line medical staff, expanding the coverage of prenatal care and establishing regional neonatal care networks^47, 48, 53-55^. In high-income regions, preterm birth prevention should focus on narrowing risk disparities among marginalized groups, such as non-Hispanic Black women in the United State^56-59^. Meanwhile, implement early risk screening and intensive prenatal care for high-risk populations^60^.

Different intervention measures should be adopted for different age groups. For infants aged 0-6 days, priority should be given to neonatal resuscitation, treating acute complications, and promoting kangaroo mother care (KMC)^61-65^. As a low-cost and high-efficiency intervention measure, KMC has been proven to effectively reduce the death of preterm infants in low-resource settings^65-67^. For infants aged 6-11 months, early developmental screening, nutritional support, and management of chronic complications can reduce long-term disability and improve quality of life^68-73^. For children aged 1-4 years, long-term follow-up and early intervention for neurodevelopmental delay can reduce the lifelong impact of preterm birth^74, 75^.

This study has several limitations. Firstly, as a retrospective secondary data analysis based on the GBD 2023 database, it can only explore the associations between variables but cannot confirm causality. Secondly, preterm birth surveillance data are incomplete in some low-income regions such as Sub-Saharan Africa, which may bias regional disease burden estimates. Thirdly, long-term projections of preterm birth burden up to 2050 do not account for unforeseeable factors including public health emergencies and major social policy adjustments.

The 0-6 days acute care and 6-11 months postnatal follow-up identified in this study may be adopted as standard clinical practice for preterm birth management, allowing for standardized diagnosis and treatment. Screening for PM_2.5_ exposure, micronutrient status, and socioeconomic risks should be incorporated into routine prenatal and neonatal care, with targeted interventions provided for high-risk individuals. Wide implementation of KMC, breast milk fortification, and regular micronutrient supplementation across all regions-particularly in low-resource settings-can effectively improve the prognosis of preterm infants at a relatively low cost.

## Conclusions

Although the global health burden of preterm birth has declined in recent decades, its rising prevalence remains a major public health challenge. Based on the GBD 2023 database, this study provides a comprehensive analysis of the global preterm birth burden from 1990 to 2023 and projections to 2050, with stratified analyses by SDI level, income group, and country. Over the past 33 years, global preterm birth-related death and DALYs have decreased significantly due to maternal and neonatal health interventions, but prevalence has continued to increase, with substantial regional and age-related disparities. Low SDI regions and lower-middle-income countries carry the heaviest disease burden. This study identifies critical intervention windows and modifiable risk factors, offering evidence for targeted global preterm birth strategies. Targeted prevention and control measures are urgently needed to reduce the global burden. Achieving the UN SDG target of ending preventable newborn deaths by 2030 requires multi-level and cross-sectoral collaboration, including strengthening health systems in high-burden regions, addressing modifiable risk factors, implementing age-specific interventions, and prioritizing preterm birth control in global health and climate agendas. Addressing persistent regional inequalities and rising prevalence will ensure equitable access to quality care for all preterm infants and advance fair development in maternal and neonatal health services.

## Data Availability

All relevant data are within the manuscript and its Supporting Information files.

http://ghdx.healthdata.org/gbd-results-tool

## Declarations

### Ethics approval and consent to participate

Not applicable.

### Consent for publication

Not applicable.

### Availability of data and materials

The datasets generated and/or analysed during the current study are available in the GBD repository, http://ghdx.healthdata.org/gbd-results-tool.

### Author contributions: CRediT

HW: Conceptualization, Data curation, Formal analysis, Writing - Original Draft. XZ: Investigation, Validation, Writing - Review & Editing. XZ: Supervision, Project administration, Writing - Review & Editing.

## Acknowledgements

Not applicable.

